# Genetic architecture of Alzheimer’s disease-related plasma biomarkers

**DOI:** 10.64898/2025.12.17.25342458

**Authors:** A Aaltonen, T Palviainen, S Heikkinen, SK Herukka, M Hiltunen, T Kokkola, S Kärkkäinen, FinnGen, A Palotie, H Runz, V Julkunen, J Kaprio, TT Saari, E Vuoksimaa

**Author notes:** Corresponding author: Aino Aaltonen.

## Abstract

**INTRODUCTION:** Alzheimer’s disease-related plasma biomarker interrelationships and factors underlying these associations remain poorly understood. To address this, we studied the heritability of plasma biomarkers and their associations with cognition.

**METHODS:** This study included 696 twins, aged 65–85, without a diagnosis of Alzheimer’s disease. Plasma amyloid beta 42 and 40, phosphorylated tau (p-tau) 181 and 217, neurofilament light chain (NfL) and glial fibrillary acidic protein (GFAP) were quantified with Simoa HD-X, and cognition was assessed with cCOG. Statistical analyses were done with multivariate twin models and linear mixed effect models.

**RESULTS:** Phenotypic associations between p-tau181, p-tau217, NfL and GFAP were 0.22–0.73. Heritability estimates were 0.39–0.66, and genetic correlations were 0.32–0.67. P-tau181 and p-tau217 were associated with cognition (β = −0.12; 95% CI: −0.19–-0.04 and β = −0.19; 95% CI: −0.26–-0.12, respectively).

**DISCUSSION:** The observed phenotypic and genetic associations highlight shared underlying mechanisms among plasma biomarkers.

## 1. Background

Alzheimer’s disease (AD) is a progressive neurological condition and the most common cause of dementia [1]. It is characterized by accumulation of extracellular amyloid beta plaques and intracellular neurofibrillary tangles, which are aggregates of hyperphosphorylated tau [2]. Neuropathological changes cause synaptic loss and neurodegeneration. The clinical picture of AD is varied, but amnestic problems are the most common early symptoms [1]. Neuropathological changes can start already in middle age [3,4], and cognitive and functional decline have been observed 6–8 years before diagnosis [4,5]. Studying individuals without diagnosis of dementia is therefore crucial for the development of early identification and interventions.

Plasma biomarkers offer a scalable and affordable way to assess pathological changes. Most promising disease-specific plasma biomarker candidates include amyloid beta 42 (Aβ42) to amyloid beta 40 (Aβ40) ratio, phosphorylated tau 181 (p-tau181) and phosphorylated tau 217 (p-tau217) [6]. Further, neurofilament light chain (NfL) and glial fibrillary acidic protein (GFAP) and have been suggested as progression markers of AD reflecting neurodegeneration and neuroinflammation, respectively. [7–10]. Plasma p-tau isoforms, especially p-tau217, have shown high accuracies for predicting amyloid positron emission tomography (PET) status, whereas Aβ42/Aβ40, NfL and GFAP have lower discriminative performance [11].

Plasma biomarkers have been investigated in relation to other biomarkers, such as amyloid PET, but their interrelationships and the factors underlying these associations have received less attention. Family designs allow disentangling genetic and environmental contributions to phenotypic variance, and a multivariate approach enables decomposition of phenotypic correlations into genetic and environmental components. Despite the relevance of phosphorylated tau in AD, only one previous study estimating the multivariate heritability of AD-related plasma biomarkers has included p-tau217, along with our previous study on the univariate heritability of p-tau217 [12,13]. These studies yielded heritability estimates of 0.56 and 0.41–0.42 [12,13]. To our knowledge, no previous family studies have included multiple isoforms of p-tau.

Investigating AD-related plasma biomarkers in relation to cognition in people without dementia offers insight into mechanisms relevant to the early identification of cognitive impairment. Previously, plasma p-tau217 has shown a stronger association with memory than other p-tau isoforms or other AD-related plasma biomarkers [14,15]. Notably, the strength of the association has been higher when participants with mild cognitive impairment (MCI) or dementia due to AD have been included [14,15], highlighting the importance of population-based research.

Here, we conducted a cross-sectional study in a population-based cohort of 65–85-year-old twins without a diagnosis of AD or other neurodegenerative condition known to affect cognition. The first aim was to assess the phenotypic associations between plasma biomarkers Aβ42/Aβ40, p-tau181, p-tau217, NfL and GFAP. The second aim was to assess genetic and environmental contributions to plasma biomarkers and their covariance. The third aim was to assess phenotypic associations between plasma biomarkers and cognition.

## 2. Methods

### 2.1. Study design

Participants of this study are twins from a cross-sectional, population-based TWINGEN study [16]. The data were gathered at six study centers in Finland: The Institute for Molecular Medicine Finland of University of Helsinki, the clinical laboratory of Turku University of Applied Sciences, Central Finland Biobank (Jyväskylä), Biobank of Eastern Finland (Kuopio), Arctic Biobank and Biobank Borealis of Northern Finland (Oulu) and Finnish Clinical Biobank Tampere [16]. During a research visit, participants completed computerized cognitive assessment and donated a blood sample for biomarker assessment. Data collection took place between February 2023 and November 2023.

### 2.2. Participants

The TWINGEN study included participants from the older Finnish Twin Cohort study who were also participants of THL Biobank [17]. Participants were 65–86 years old, and they were included irrespective of their co-twin’s participation. They were of European ancestry and spoke Finnish as their first language. Exclusion criteria included a diagnosis of AD or another dementia-causing neurodegenerative disease, major cognition-affecting neurological or developmental condition and a serious psychiatric condition based on health registry information [16]. The exclusion criteria were applied to 2022 health registry data and eligibility was further confirmed through a telephone call before participation.

*APOE* genotype was determined from genome-wide genotyping data [18]. *APOE* ε4 status was considered positive for participants with at least one ε4 allele. Information on participants’ sex and date of birth was retrieved from a population registry. Lifetime years of education were transformed from self-reported education level (see Aaltonen et al. [19] for details). For further information about the TWINGEN cohort, see the published study protocol [16].

### 2.3. Biomarker assessment

Biomarker analyses were conducted at the Biomarker Laboratory of University of Eastern Finland. We obtained the following plasma biomarkers: p-tau181, p-tau217, Aβ42, Aβ40, GFAP and NfL; all quantified with Simoa HD-X Analyzer (Quanterix, Billerica, Massachusetts, USA). We measured p-tau181 with Advantage V2.1 Kit (Ref# 104111, Quanterix), p-tau217 with ALZpath Simoa pTau-217 v2 Assay Kit (Ref# 104371, Quanterix) and other biomarkers with Simoa Neurology 4-Plex E Advantage Kit (Ref# 103670, Quanterix). The EDTA plasma samples were thawed, mixed and centrifuged (10 000 × g, 5 min, +20°C) prior to the analyses.

### 2.4. Cognitive assessment

Cognition was assessed with a web-based tool developed for detection of MCI and dementia due to neurodegenerative diseases called cCOG [19,20]. We used the total cognitive score, which had a range of 0–1, with greater values indicating better cognitive functioning. The total score was calculated as a weighted sum from the number of correctly recalled words in Episodic Memory Task (12-word list with three trials of learning and a delayed free recall) and time in Modified Trail Making A and B Tasks (TMT-A and TMT-B). Outliers were clipped to the score range edges.

### 2.5. Twin design

We assessed the phenotypic associations and genetic and environmental contributions and correlations between the biomarkers with a multivariate AC/DE model. AC/DE models are structural equation models that estimate phenotypic variance and decompose it into additive genetic (A), common environmental (C) or non-additive genetic (D) and unique environmental (E) variance components by using monozygotic (MZ) and dizygotic (DZ) twin pairs [21]. Additive genetic effects reflect the combined impact of individual alleles, and as MZ twins are genetically (nearly) identical and DZ twins share on average 50% of their segregating genes, the additive genetic correlation within twin pairs is fixed at r_A_ = 1 for MZ twins and at r_A_ = 0.5 for DZ twins. C effects are environmental effects that make co-twins similar, and the shared environmental correlation is fixed at r_C_ = 1 across all twin pairs. D effects reflect dominance, which refers to interaction between two alleles within a locus, and epistasis, which means interaction between two loci. Thus, the non-additive genetic correlation is fixed at r_D_ = 1 for MZ twins and r_D_ = 0.25 for DZ twins. E effects are environmental effects unique to each twin and uncorrelated by definition. They also capture measurement error.

As twin data does not allow simultaneous estimation of A, C and D effects, we elected to use an ACE model, where D effects are fixed to zero. In ACE models, expected phenotypic covariance in MZ pairs can be expressed as Cov(P_Twin1_, P_Twin2_) = A + C and in DZ pairs as Cov(P_Twin1_, P_Twin2_) = 0.5 × A + C, while expected variance is expressed as Var(P) = A + C + E. Taking advantage of the different expected covariance structures for MZ and DZ pairs allows for the estimation of the A, C and E variance components.

The multivariate ACE model extends the univariate model by including multiple phenotypes in the model, and phenotypic variance-covariance structure is decomposed to A, C and E components. In our analysis, we used freely estimated symmetrical matrices to estimate A, C and E variance-covariance matrices.

### 2.6. Statistical methods

#### 2.6.1. Genetic and environmental effects on plasma biomarkers

Neither the raw biomarker values regressed on age and sex, nor log-transformed biomarker values regressed on age and sex were normally distributed across all biomarkers. Further, log-transformed residuals did not have equal variances across randomized twin order. Therefore, we used the Blom transformation (c = 3/8) to obtain rank-based inverse normalized biomarker values and regressed them on age and sex. The standardized residuals were used in the twin models.

Prior to model fitting, we calculated the phenotypic intraclass correlations (ICC) for MZ and DZ twin pairs with standardized residuals of rank-based inverse normalized biomarkers with irr package [22]. For p-tau181, NfL and GFAP, the ICCs were consistent with r_MZ_ < 2 × r_DZ_, whereas p-tau217 showed r_MZ_ > 2 × r_DZ_. On contrast, Aβ42/Aβ40 had higher correlations for DZ than for MZ pairs and was therefore excluded from the models. We did not include Aβ42 and Aβ40 individually in the models, as their separate consideration is less biologically meaningful, and Aβ42 had only marginally larger ICC for MZ pairs than for DZ pairs.

Based on the ICCs, we opted to fit C effects instead of D effects. First, we compared the multivariate ACE model to a saturated model with means, variances and covariances freely estimated. Then, we fitted AE, CE and E sub-models by fixing the 1) C, 2) A and 3) C and A variance matrices to 0 and compared each sub-model to the full ACE model with likelihood ratio tests and Akaike information criteria (AIC). We report the phenotypic correlations, standardized variance components and variance component correlations for the best fitting model. Additionally, we report standardized variance components for the full ACE model and phenotypic full and partial Spearman correlation for raw biomarker values in supplementary materials.

The twin models were fitted with OpenMx package [23], and full information maximum likelihood (FIML) was used for model estimation. Variance–covariance matrices were estimated freely. We acknowledge that this approach may result in negative variance or covariance estimates, however, it maintains correct Type I/II error rates and avoids biasing the estimates that would be introduced by restricting the estimates to be positive [24].

#### 2.6.2. Association between plasma biomarker levels and cognitive functioning

The associations between plasma biomarker levels and cognitive functioning were analyzed with linear mixed-effects models using lme4 package [25]. Total cognitive score was used as the outcome variable. First, we fitted a covariate model with age, sex and lifetime years of education as fixed effects, and study site and family effects as random intercepts. Next, we fitted a separate model for each investigated biomarker. In addition to the covariates included in the covariate model, these models included 1) log-ratio and geometric mean in the log scale of Aβ42 and Aβ40, 2) p-tau181, 3) p-tau217, 4) NfL, 5) GFAP and 6) all plasma biomarkers as fixed effects. The log-ratio log(Aβ42) - log(Aβ40) was chosen due to the multiplicative nature of the ratio variable, and the geometric mean 0.5(log(Aβ42) + log(Aβ40)) was included to account for total accumulation of amyloid beta. Continuous fixed effects and the outcome variable were standardized before model fitting.

We assessed whether inclusion of a given plasma biomarker resulted in improved model fit by evaluating likelihood ratio against a χ^2^-distribution and corrected for multiple testing by adjusting the p-values with the Benjamini–Yekutieli procedure to control the false discovery rate. The marginal R^2^ values, representing variance explained by the fixed effects, were obtained according to Nakagawa and Schielzeth [26] and calculated with MuMIn package [27].

### 2.7. Statistical considerations

Statistical significance was set at α = 0.05. Analyses were done with R statistical software (version 4.5.0) [28].

Some data were missing for 60 (8.62%) participants (Table S1). For the multivariate ACE analyses, missing biomarker levels were left as missing and imputed median value of 10 years of education was used as a covariate. When calculating Spearman correlation coefficients, we used pairwise deletion and imputed education value for the partial correlation. For linear mixed-effects models, participants with any missing biomarker level or cCOG total score were excluded and imputed median value for missing education level was used. *APOE* ε4 status is reported in descriptive statistics, but we excluded it as a covariate to enhance clinical interpretability.

Analysis code is available at https://osf.io/hk6f9.

## 3. Results

### 3.1. Descriptive statistics

There were 2718 individuals invited to the TWINGEN study, and 704 participated. Out of them, 8 participants did not donate a blood sample and were excluded from the current study, resulting in a sample size of 696, including 153 full twin pairs (80 MZ and 73 DZ). Of the 80 MZ twin pairs, 56 were female pairs and 24 were male pairs. Of the 73 DZ pairs, 33 were female pairs, 20 were male pairs and 20 were male-female pairs.

Demographic characteristics of the participants are presented in Table 1. Most participants (n = 651) completed the cCOG during the study visit, however, 26 participants did it at home due to issues with login credentials.

**Table 1.** Demographic characteristics of the study sample.

| Variable | All | Sex |  | Co-twin |  |
| --- | --- | --- | --- | --- | --- |
|  |  | Male | Female | Data from both co-twins | Data from co-twin missing |
| Number of participants | 696 | 298 | 398 | 390 | 306 |
| Female | 398 (57.18) | 0 (0.00) | 398 (100.00) | 200 (51.28) | 198 (64.71) |
| *APOE ε4 | 203 (29.21) | 87 (29.29) | 116 (29.15) | 110 (28.28) | 93 (30.39) |
| Age, yr | 76.17 (4.57) | 76.64 (4.38) | 75.81 (4.68) | 77.33 (3.97) | 74.68 (4.85) |
| Aβ40, pg/ml | 114.45 (31.79) | 113.01 (31.36) | 115.52 (32.10) | 114.44 (31.39) | 114.46 (32.33) |
| Aβ42, pg/ml | 6.56 (2.07) | 6.26 (2.00) | 6.79 (2.10) | 6.55 (1.99) | 6.58 (2.17) |
| Aβ42/Aβ40 | 0.06 (0.03) | 0.06 (0.02) | 0.06 (0.03) | 0.06 (0.02) | 0.06 (0.03) |
| p-tau181, pg/ml | 27.31 (13.01) | 29.43 (15.02) | 25.72 (11.03) | 27.89 (12.92) | 26.56 (13.11) |
| p-tau217, pg/ml | 0.45 (0.31) | 0.47 (0.32) | 0.43 (0.31) | 0.45 (0.30) | 0.44 (0.33) |
| NfL, pg/ml | 24.69 (14.12) | 24.55 (11.65) | 24.80 (15.74) | 24.95 (13.52) | 24.35 (14.87) |
| GFAP, pg/ml | 156.98 (85.23) | 136.39 (59.92) | 172.44 (97.37) | 161.71 (98.24) | 150.97 (64.72) |
| cCOG WL immediate,<br>words | 17.83 (6.07) | 17.06 (5.74) | 18.42 (6.25) | 17.57 (6.38) | 18.16 (5.65) |
| cCOG WL delayed,<br>words | 6.51 (2.43) | 6.20 (2.36) | 6.74 (2.46) | 6.27 (2.48) | 6.81 (2.34) |
| cCOG TMT-A, s | 75.58 (48.91) | 76.31 (53.34) | 75.03 (45.32) | 77.17 (50.61) | 73.57 (46.68) |
| cCOG TMT-B, s | 251.57 (124.13) | 259.96 (131.23) | 245.24 (118.29) | 252.11 (126.35) | 250.89 (121.50) |
| cCOG total score | 0.60 (0.16) | 0.58 (0.16) | 0.61 (0.16) | 0.59 (0.17) | 0.61 (0.14) |

Abbreviations: Aβ40, amyloid beta 40; Aβ42, amyloid beta 42; GFAP, glial fibrillary acidic protein; NfL, neurofilament light chain; p-tau181, phosphorylated tau 181; p-tau217, phosphorylated tau 217; TMT; trail making test; WL, word list task.
Note: Means (standard deviations) are presented for continuous variables, and number of participants (proportions) are presented for categorical variables (Female and *APOE* ε4). *APOE* ε4 represents the number of participants with at least one ε4 allele. Data from both co-twins describes individuals who participated with their co-twin, and data from co-twin missing is describes those whose co-twin did not participate.
\**APOE* genotype was missing for 1 participant.

### 3.2. Twin models

Genetic and environmental effects on plasma biomarkers were assessed with multivariate twin models. Prior to fitting the ACE model, we calculated phenotypic ICCs (Table 2) and fitted a saturated model with means, variances and covariances freely estimated. We then tested the assumptions of equal means, variances and covariances across twin order, and equal means and variances across zygosity. The constrained model did not fit significantly worse than the saturated model (χ^2^_36_ = 39.73; p = 0.3073) and had lower AIC (ΔAIC = −32.27), indicating that the assumptions were met. We then proceeded to fit the ACE model and tested it against the saturated model. Again, the model fit was not significantly worse compared to the saturated model (χ^2^_42_ = 46.06; p = 0.3079; ΔAIC = −37.94). The standardized variance component estimates from the ACE model are presented in Table S3.

**Table 2.**
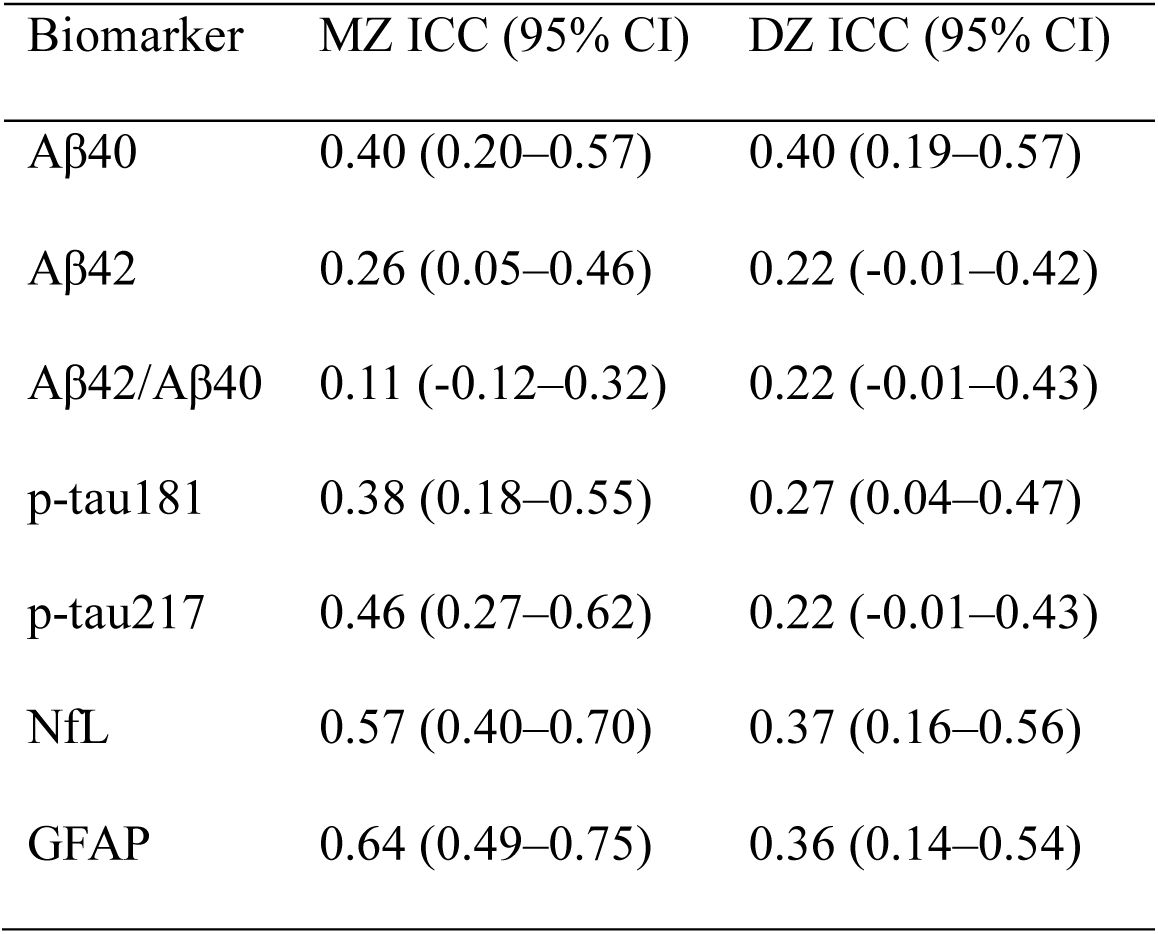
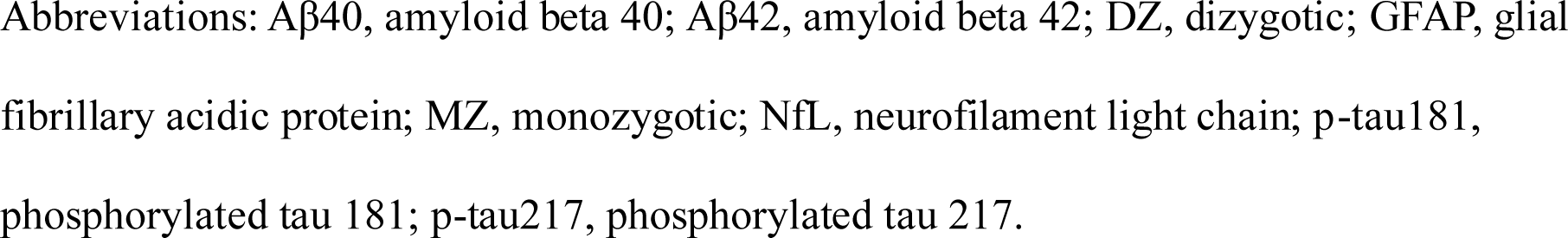
Phenotypic intraclass correlation estimates and 95% confidence intervals for standardized residuals of rank-based inverse normalized biomarkers for MZ and DZ twin pairs.

| Biomarker | MZ ICC (95% CI) | DZ ICC (95% CI) |
| --- | --- | --- |
| Aβ40 | 0.40 (0.20–0.57) | 0.40 (0.19–0.57) |
| Aβ42 | 0.26 (0.05–0.46) | 0.22 (-0.01–0.42) |
| Aβ42/Aβ40 | 0.11 (-0.12–0.32) | 0.22 (-0.01–0.43) |
| p-tau181 | 0.38 (0.18–0.55) | 0.27 (0.04–0.47) |
| p-tau217 | 0.46 (0.27–0.62) | 0.22 (-0.01–0.43) |
| NfL | 0.57 (0.40–0.70) | 0.37 (0.16–0.56) |
| GFAP | 0.64 (0.49–0.75) | 0.36 (0.14–0.54) |

Abbreviations: Aβ40, amyloid beta 40; Aβ42, amyloid beta 42; DZ, dizygotic; GFAP, glial fibrillary acidic protein; MZ, monozygotic; NfL, neurofilament light chain; p-tau181, phosphorylated tau 181; p-tau217, phosphorylated tau 217.

Next, we fit the AE, CE and E sub-models. The AE model did not show statistically significant worsening of fit compared to the ACE model, unlike the CE and E sub-models, that fit significantly worse (Table S2). Further, the AE model had the lowest AIC-value (Table S2). Subsequently, the AE model was chosen as the best fitting model (Figure 1).

**Figure 1.**
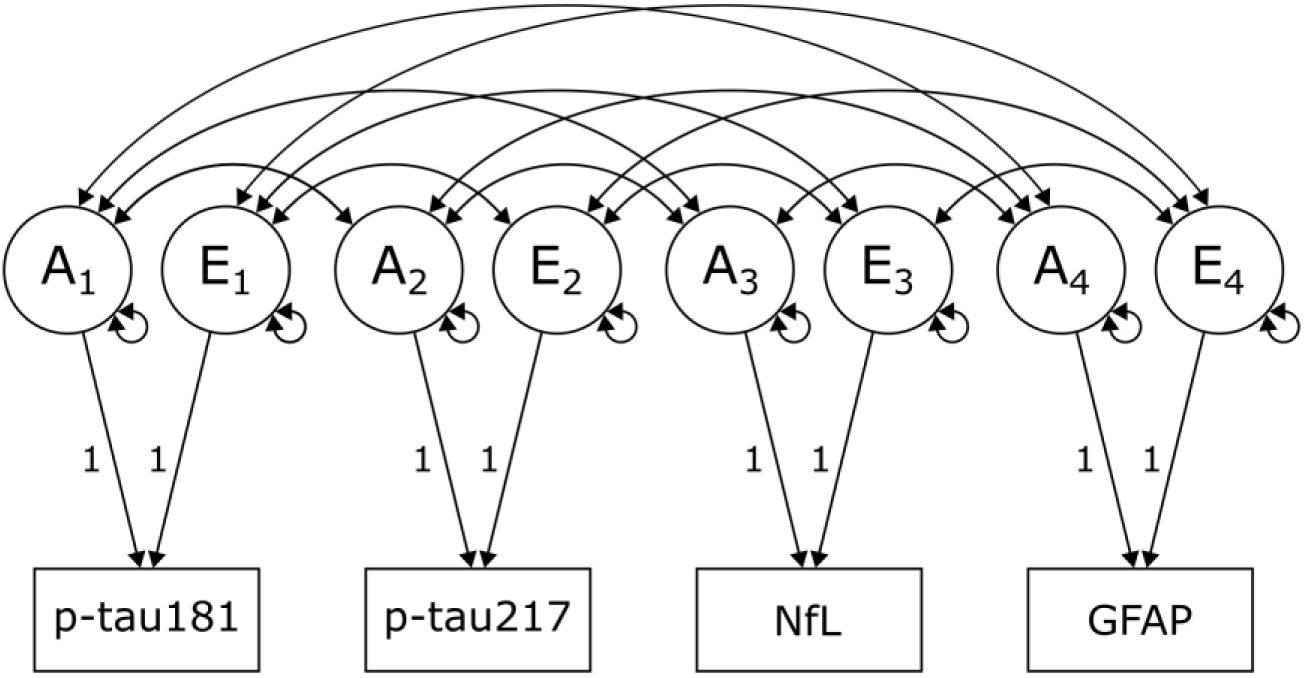
Multivariate ACE model, with variance of p-tau181, p-tau217, NfL and GFAP decomposed into additive genetic (A_1_-A_4_) and unique environmental (E_1_-E_4_) components. Abbreviations: GFAP, glial fibrillary acidic protein; NfL, neurofilament light chain; p-tau181, phosphorylated tau 181; p-tau217, phosphorylated tau 217.

The phenotypic associations are presented in Figure 2. The standardized A component estimates, reflecting heritability, varied between 0.39 (p-tau181) and 0.66 (GFAP; Table 3). The correlation estimates between the A components and the estimates on how much the genetic correlation explains of the phenotypic correlation are presented in Figure 3, and the correlation estimates between the E components are presented in Figure 4.

**Figure 2.**
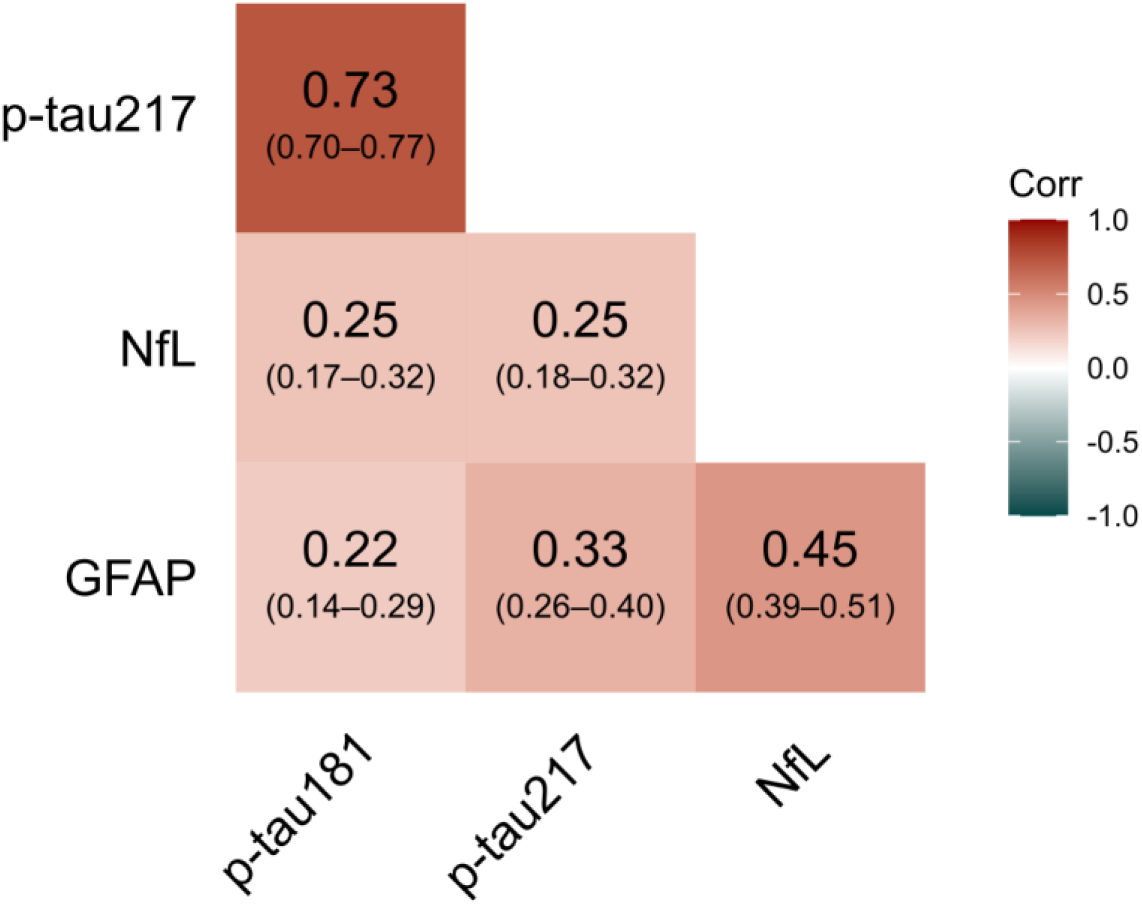
Model-implied phenotypic pairwise correlation estimates with 95% confidence intervals from the fitted AE model. Abbreviations: GFAP, glial fibrillary acidic protein; Corr, correlation coefficient; NfL, neurofilament light chain; p-tau181, phosphorylated tau 181; p-tau217, phosphorylated tau 217.

**Figure 3.**
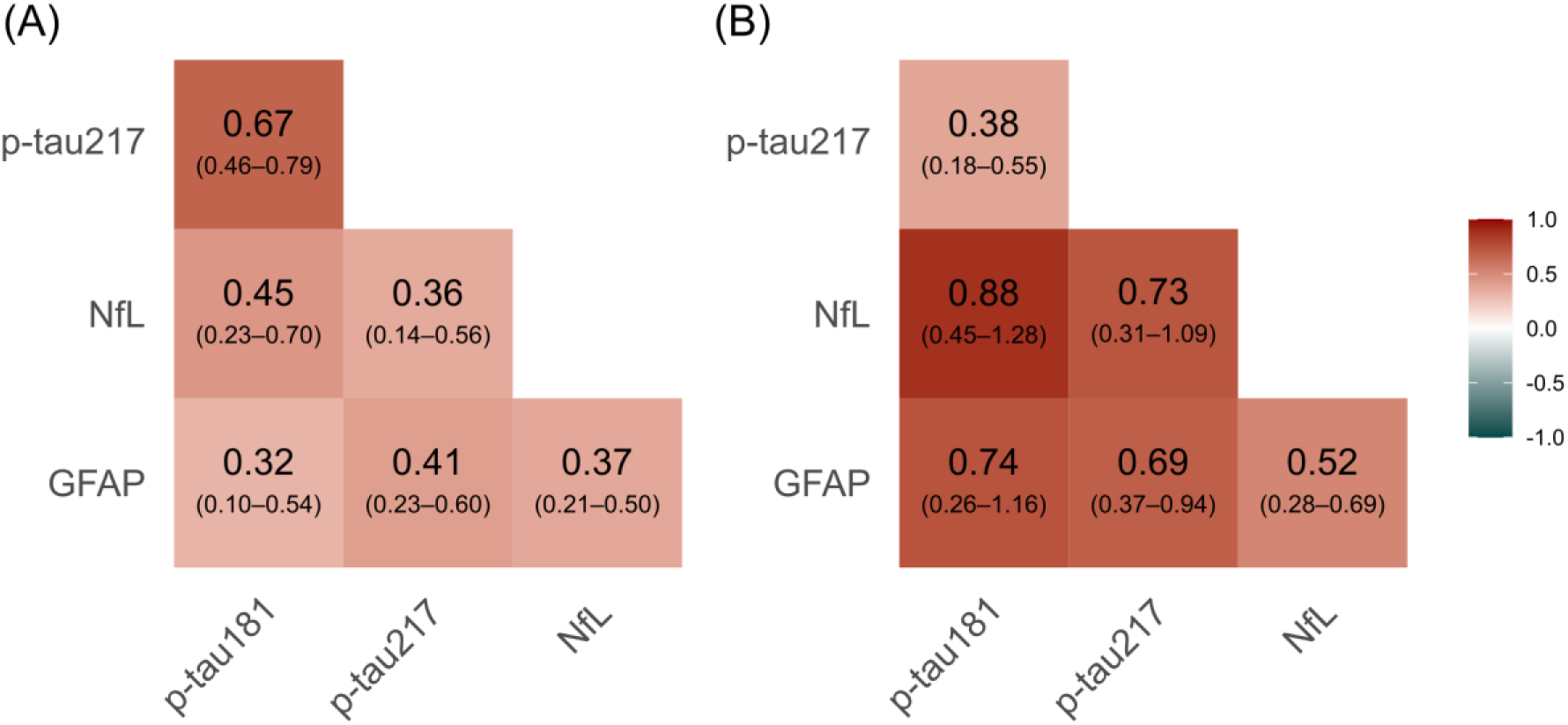
Model-implied estimates with 95% confidence intervals from the fitted AE model for (A) pairwise correlations between the latent additive genetic components, and (B) proportions of covariance explained by the genetic correlations. Abbreviations: GFAP, glial fibrillary acidic protein; Corr, correlation coefficient; NfL, neurofilament light chain; p-tau181, phosphorylated tau 181; p-tau217, phosphorylated tau 217.

**Figure 4.**
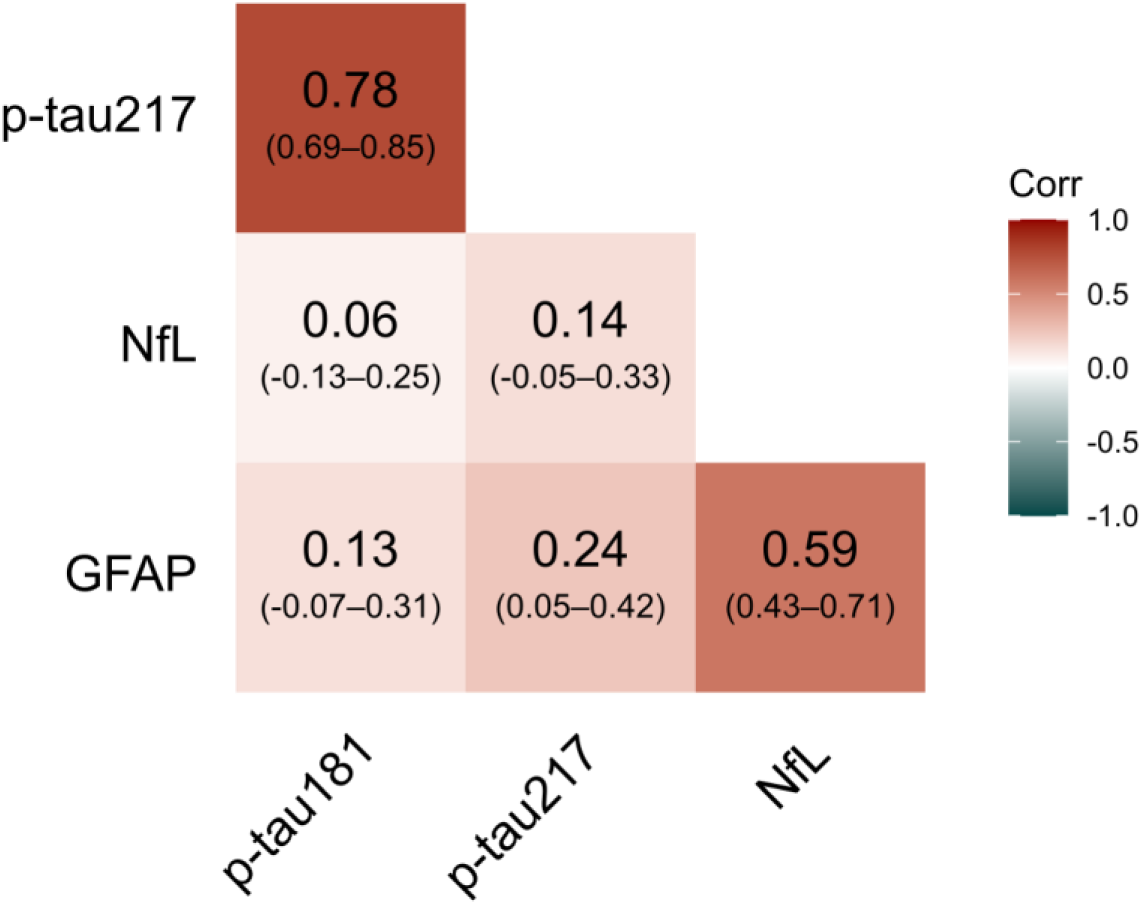
Model-implied pairwise correlation estimates with 95% confidence intervals between the latent unique environmental components from the fitted AE model. Abbreviations: GFAP, glial fibrillary acidic protein; NfL, neurofilament light chain; p-tau181, phosphorylated tau 181; p-tau217, phosphorylated tau 217.

**Table 3.**
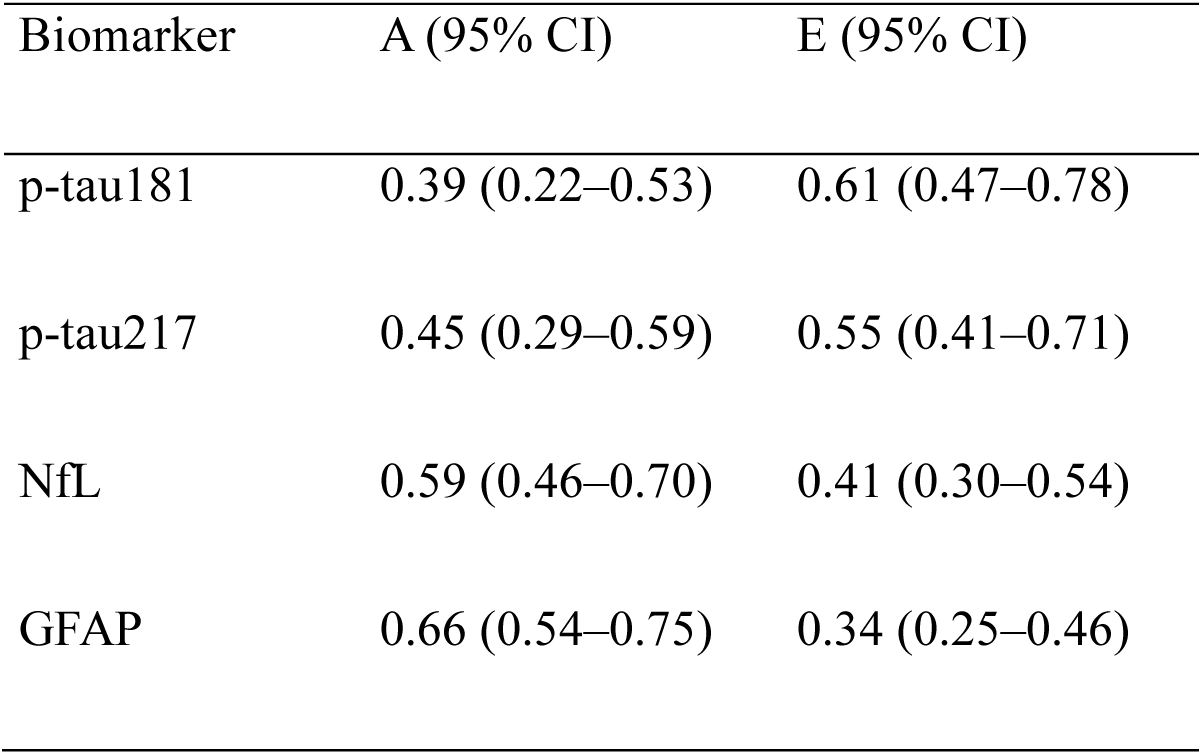
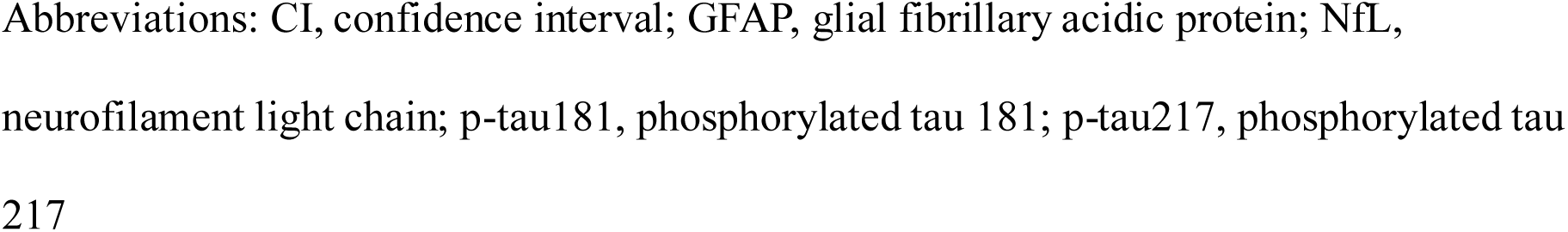
Standardized estimates for additive genetic (A) and unique environmental (E) variance components with 95% confidence intervals from the fitted AE model.

| Biomarker | A (95% CI) | E (95% CI) |
| --- | --- | --- |
| p-tau181 | 0.39 (0.22–0.53) | 0.61 (0.47–0.78) |
| p-tau217 | 0.45 (0.29–0.59) | 0.55 (0.41–0.71) |
| NfL | 0.59 (0.46–0.70) | 0.41 (0.30–0.54) |
| GFAP | 0.66 (0.54–0.75) | 0.34 (0.25–0.46) |

Abbreviations: CI, confidence interval; GFAP, glial fibrillary acidic protein; NfL, neurofilament light chain; p-tau181, phosphorylated tau 181; p-tau217, phosphorylated tau 217

Partial Spearman correlations between the biomarker values are presented in Figures S1-S2, and the variance component estimates in the full ACE model are presented in Table S2.

### 3.3. Associations between plasma biomarkers and cognitive functioning

Associations between the plasma biomarkers and cognition were analyzed with linear mixed-effects models. In the nested covariate model, female sex and longer education were positively associated with cognition (β = 0.18, 95% confidence interval (CI): 0.04–0.32 and β = 0.27, 95% CI: 0.20–0.34), whereas age had negative association with cognition (β = −0.30, 95% CI: −0.37–-0.22). Model 2 (p-tau181), Model 3 (p-tau217) and Model 6 (all biomarkers) had statistically significantly better model fit than model with covariates alone, while Models 4–6 did not (Table S4). The standardized beta coefficient was β = −0.12 (95% CI: −0.19–-0.04) for p-tau181 in Model 2 and β = −0.19 (95% CI: −0.26–-0.12) for p-tau217 in Model 3 (Figure 5).

**Figure 5.**
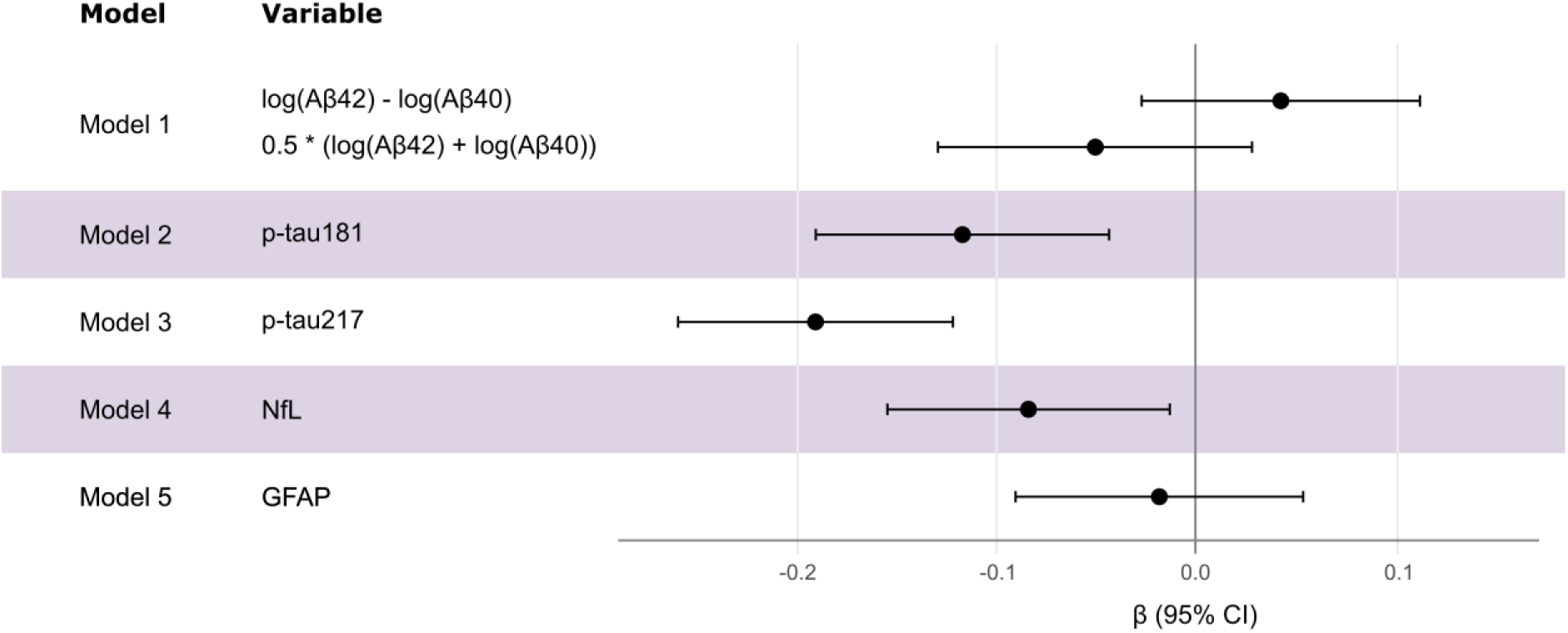
Standardized beta coefficients with 95% confidence intervals for plasma biomarkers in linear mixed effects models 1–5. All models included sex, education and age as fixed effects, study site and family effects as random intercepts and total cognitive score as the outcome. Abbreviations: Aβ40, amyloid beta 40; Aβ42, amyloid beta 42; CI, confidence interval; GFAP, glial fibrillary acidic protein; NfL, neurofilament light chain; p-tau181, phosphorylated tau 181; p-tau217, phosphorylated tau 217.

## 4. Discussion

We investigated 1) the phenotypic associations between plasma biomarkers of AD, 2) the genetic and environmental contributions to plasma biomarkers and their covariance, and 3) the phenotypic associations between plasma biomarkers and cognition.

We observed a strong phenotypic correlation between p-tau181 and p-tau217, consistent with findings by Mielke et al. [29]. While p-tau217 is more accurate for capturing AD-related pathological changes, both are strongly associated with AD-pathology [30], and their phenotypic association was expected.

Both p-tau isoforms had weak but statistically significant phenotypic associations with NfL and GFAP. NfL is used to characterize neuronal injury and is not specific to Alzheimer’s disease [7]. The rate of change in NfL corresponds with the rate of change in amyloid PET [31]. However, the cross-sectional association between plasma NfL and cortical atrophy has been observed in participants with MCI or AD, but not with participants without cognitive impairment or Aβ load [9], indicating that NfL reflects neurodegenerative processes more clearly in the presence of AD-related pathology. GFAP reflects astrocytic activation, and in AD, astrocytes respond to Aβ plaque deposition and may contribute to overall Aβ accumulation [32]. Longitudinal changes in plasma GFAP correspond closely with changes in amyloid PET [31], and plasma GFAP has been linked to Aβ-PET measures throughout the disease continuum [33]. In contrast, its association with tau-PET emerges only in middle and late Braak stages and is attenuated when Aβ-PET is accounted for [33], potentially explaining the weaker association with p-tau isoforms in participants without diagnosis of dementia or AD.

Finally, we found a moderate phenotypic association between NfL and GFAP. Given the bidirectional relationship between neurodegeneration and neuroinflammation [34], the phenotypic association is not surprising.

In line with previous studies, we found that additive genetic effects and unique environmental factors contributed to the phenotypic variance-covariance structure, while common environmental factors had negligible effect [12,13,35,36]. The standardized heritability estimates ranged from 0.39 to 0.66 for p-tau181, p-tau217, NfL and GFAP.

We did not observe higher correlations for Aβ42/Aβ40 in MZ twin pairs than in DZ twin pairs, indicating zero or negligible heritability for this plasma biomarker ratio measure. Therefore, we excluded amyloid beta measures from the multivariate model. In line with this, previous studies reported zero [35] or low (0.16) [36] heritability for the Aβ42/Aβ40 ratio. [36]. Changes in plasma Aβ42/Aβ40 may occur before changes in p-tau isoforms, NfL or GFAP [31,37]. However, variance in plasma Aβ reflects peripheral Aβ in addition to AD-related brain amyloid accumulation [38], and plasma Aβ levels show smaller relative changes in the presence of cerebral Aβ and more susceptibility to variation in preanalytical handling than CSF Aβ, making this biomarker less robust than its CSF counterpart [39].

We observed several genetic and environmental correlations between the plasma biomarkers reflecting amyloid and tau pathology, neurodegeneration and neuroinflammation. All biomarkers included in the twin models had at least weak genetic correlations, implying that each biomarker pair had some overlapping genes that affect both. The highest genetic correlation (0.67) and the highest environmental correlation (0.78) were both between p-tau181 and p-tau217, implicating similar but not identical genes and environmental factors influencing their expression. Notably, only 38% of the phenotypic correlation between these isoforms was explained by genetic correlation.

In line with previous research [14,15], plasma p-tau181 and p-tau217 were associated with cognition in this cross-sectional study of participants without prior diagnosis of dementia-causing neurodegenerative diseases. However, NfL, GFAP, the log-ratio or the geometric mean in the log scale of Aβ42 and Aβ40 were not associated with cognition after controlling for multiple testing. Most previous studies targeting participants without dementia or AD pathology have observed an association between NfL and cognition [14,40–42], although conflicting results exist [43]. Our results differ slightly from a previous meta-analysis, which reported a weak association between GFAP and cognition (β = −0.09, 95% CI: −0.15–-0.03) for participants without dementia [44]. While plasma Aβ42/Aβ40 has been associated with future cognitive decline, there is no consistent cross-sectional association with cognition, in line with our findings [14,45,46].

### 4.1. Limitations and strengths

Our study has several limitations. First, the sample size for twin analyses was rather small, and we did not have adequate power to test for sex differences. These have not been previously observed for Aβ42/Aβ40, NfL or GFAP, but Rousset et al. [13] observed small sex-differences in p-tau217. Second, all participants were of European ancestry, and the results of our study may not generalize to other ethnicities.

Our study also has important strengths. We have included two p-tau isoforms in addition to Aβ42/Aβ40, NfL and GFAP, which allows for comprehensive multivariate twin analysis for core AD biomarkers and markers for disease progression. The use of a population-based cohort reduces selection bias and strengthens the external validity of our results. Lastly, excluding participants with a diagnosis of AD or dementia increases the relevance of our findings for early detection.

### 4.2. Conclusion

The heritability of plasma biomarkers p-tau181, p-tau217, NfL and GFAP ranged from 0.39 to 0.66. All biomarkers had pairwise genetic associations, and several biomarkers had environmental associations. Interestingly, while p-tau181 and p-tau217 had strong genetic and environmental correlations, neither approached unity, implying unique genes and environmental factors behind these markers, that should be considered for future genome-wide association studies and studies on risk factors. Both p-tau isoforms were associated with cognition in participants without diagnosis of dementia, highlighting their role in early identification of cognitive impairment. Overall, our results offer insight into disease-specific and progression markers of Alzheimer’s disease and inform future research examining which specific genetic and environmental factors are shared and unique to each plasma biomarker, helping uncover etiological mechanisms underlying Alzheimer’s disease pathology.

## Supporting information

FinnGen authors

## Data Availability

The TWINGEN data are stored at the THL Biobank and at FinnGen sandbox environment. The THL Biobank data are available to qualified applicants from academia and companies; for details see https://thl.fi/en/research-and-development/thl-biobank/for-researchers/application-process. The FinnGen sandbox environment is available upon reasonable request and with permission of FinnGen; for details see https://www.finngen.fi/en/researchers/accessing.

## Abbreviations

A: additive genetic effects
Aβ40: amyloid beta 40
Aβ42: amyloid beta 42
AD: Alzheimer’s disease
AIC: Akaike’s information criterion
C: common environmental effects
CI: confidence interval
D: non-additive genetic effects
DZ: dizygotic
E: unique environmental effects
GFAP: glial fibrillary acidic protein
MZ: monozygotic
NfL: neurofilament light chain
p-tau181: phosphorylated tau 181
p-tau217: phosphorylated tau 217

## Acknowledgements

The participants of the TWINGEN study were recruited through THL Biobank (study number THLBB2022_83). We thank the participants for taking part in biobank research and the TWINGEN study. We thank Mia Urjansson from University of Helsinki, Sabrina Belgasem from Helsinki Biobank, Auli Toivola from THL biobank, biobank personnel from Finnish Clinical Biobank Tampere, Arctic Biobank, Biobank Borealis of Northern Finland, Central Finland Biobank and Biobank of Eastern Finland, as well as biomedical laboratory scientist students from the clinical laboratory of Turku University of Applied Sciences for their contributions to data collection.

## Conflict of interest statement

HR is a full-time employee at Insitro Inc. AP is the Chief Scientific Officer of the FinnGen project, which is funded by industry parties listed in Funding.

## Consent statement

All participants provided written informed consent for their participation. The TWINGEN project has been reviewed and approved by the HUS Regional Committee on Medical Research Ethics (approval number 16831/2022), and research permission was granted from THL Biobank (diary ID 83/2022). The study was completed in accordance with the Declaration of Helsinki.

## Funding

This work was supported by the Finnish Cultural Foundation [grant number 00240252] and Tauno Virtanen fund to AA, the Research Council of Finland [grant number 338182], the Sigrid Jusélius Foundation, The Strategic Neuroscience Funding of the University of Eastern Finland, the Jane and Aatos Erkko Foundation and the Alzheimer’s Association [grant number ADSF-24-1284326-C] to MH, The Strategic Neuroscience Funding of the University of Eastern Finland to SH and the Sigrid Jusélius Foundation to EV. The TWINGEN study was supported by the FinnGen project that is supported by Business Finland [grant numbers HUS 4685/31/2016 and UH 4386/31/2016] and the following industry partners: AbbVie, AstraZeneca UK, Biogen, Bristol Myers Squibb (and Celgene Corporation & Celgene International II), Genentech, Merck Sharp & Dohme LLC, a subsidiary of Merck & Co., Inc., Rahway, NJ, USA, Pfizer, GlaxoSmithKline Intellectual Property Development, Sanofi US Services, Maze Therapeutics, Janssen Biotech, Novartis, and Boehringer Ingelheim. The older Finnish Twin Cohort was supported by the Research Council of Finland [grant numbers 265240, 263278 and 308248].

## Notes

### Author Declarations

The HUS Regional Committee on Medical Research Ethics gave ethical approval for this work (approval number 16831/2022).

### Summary of Updates

Supplemental files updated, minor formatting changes added.

