## Supplementary Tables and Figures for "Genetic architecture of Alzheimer’s disease-related plasma biomarkers"

### Supplement for methods

Supplementary Table 1. Variables with missing values.

| Variable | Number of missing values |
| --- | --- |
| *APOE* genotype | 1 |
| Aβ40 | 8 |
| Aβ42 | 9 |
| Aβ42/Aβ40 | 10 |
| cCOG TMT-A | 26 |
| cCOG TMT-B | 28 |
| cCOG total score | 36 |
| cCOG WL delayed | 32 |
| cCOG WL immediate | 22 |
| GFAP | 6 |
| NfL | 6 |
| Years of education | 13 |
| Zygosity | 1 |

Abbreviations: Aβ40, amyloid beta 40; Aβ42, amyloid beta 42; GFAP, glial fibrillary acidic protein; NfL, neurofilament light chain; TMT, trail making test; WL, word list task.

### Supplement for results

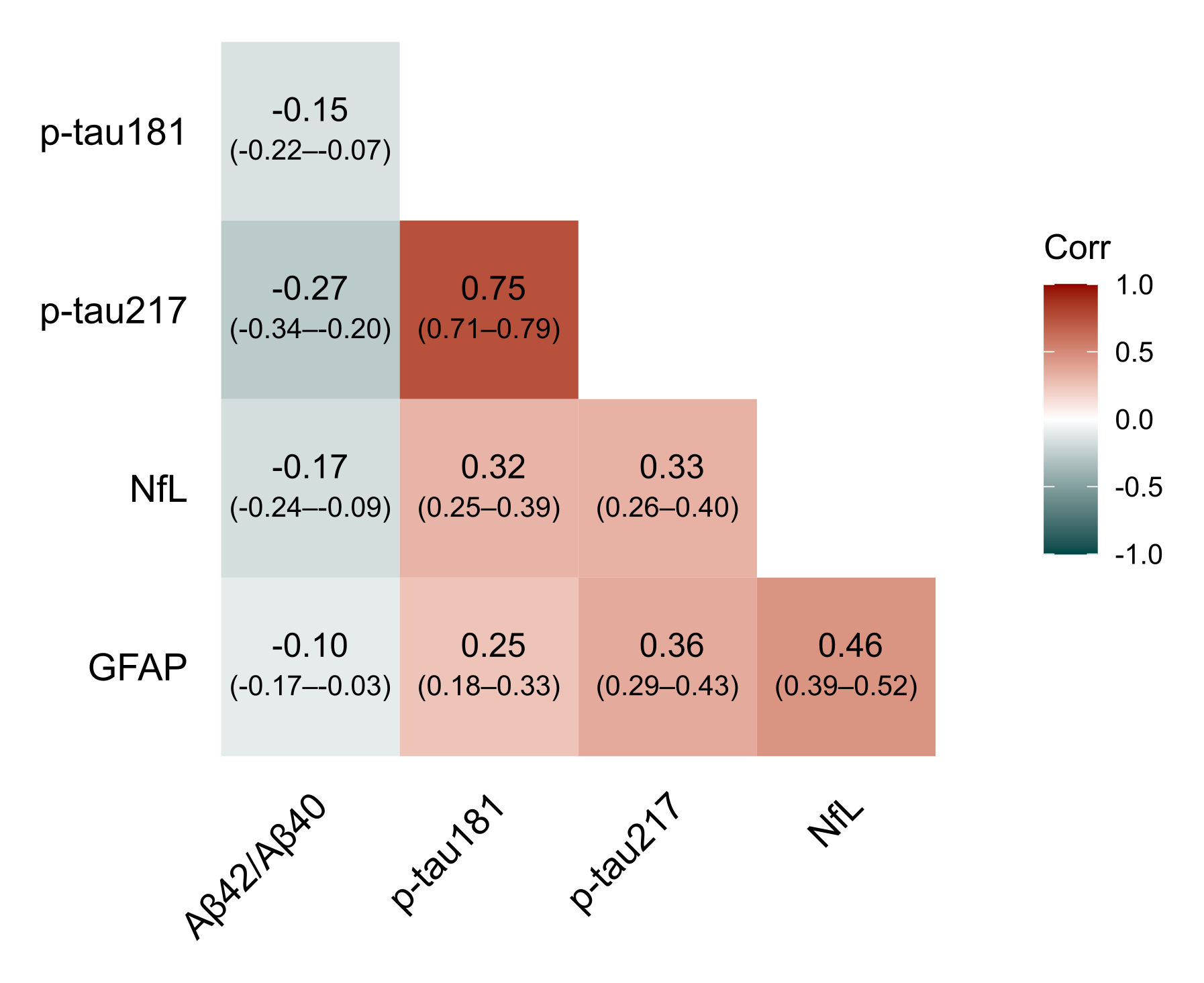

Supplementary Figure 1. Pairwise Spearman correlation coefficients with 95% confidence intervals. Abbreviations: Abbreviations: GFAP, glial fibrillary acidic protein; Corr, correlation coefficient; NfL, neurofilament light chain; p-tau181, phosphorylated tau 181; p-tau217, phosphorylated tau 217.

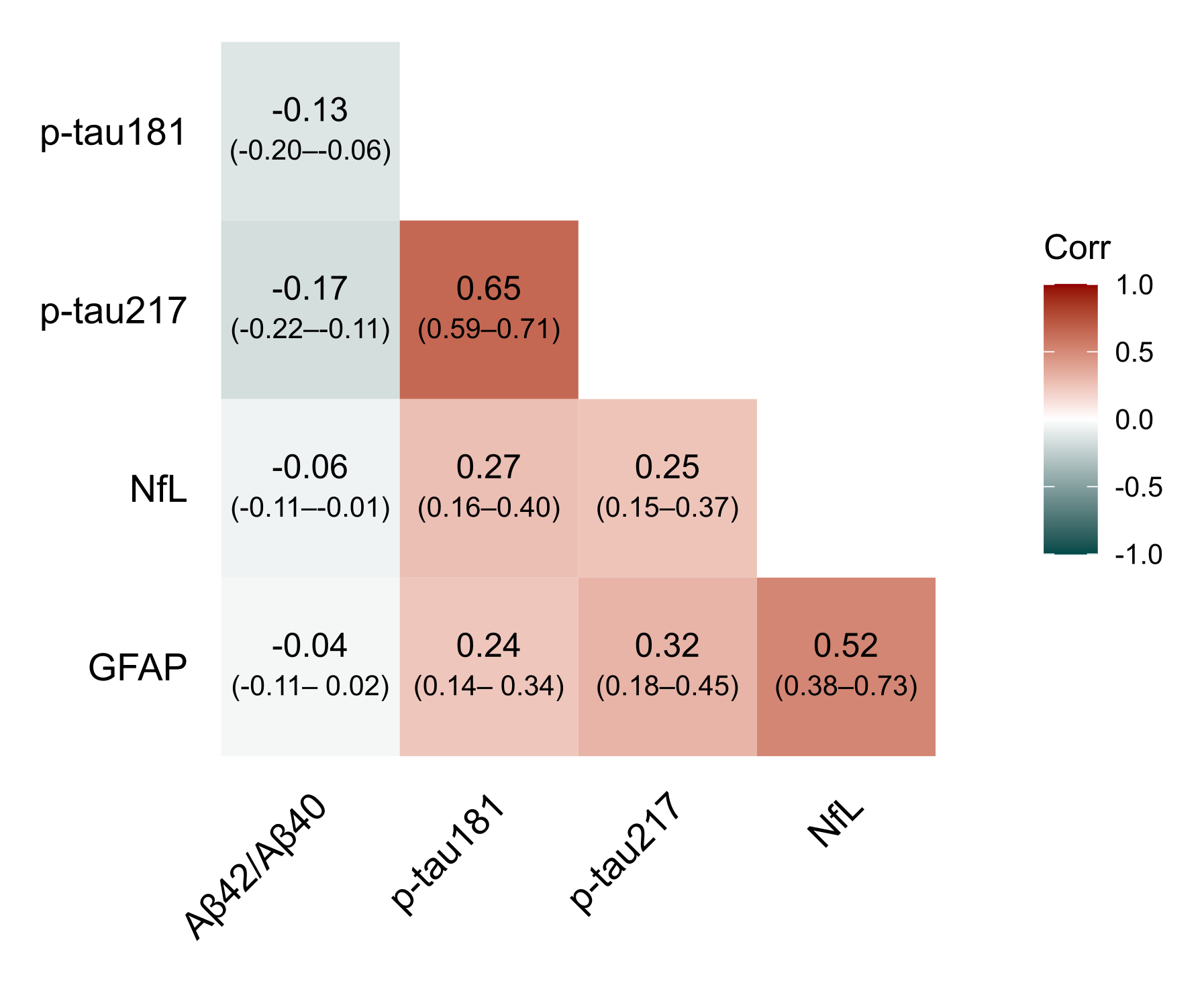

Supplementary Figure 2. Pairwise partial Spearman correlation coefficients with 95% confidence intervals, controlling for age and sex. Abbreviations: Abbreviations: GFAP, glial fibrillary acidic protein; Corr, correlation coefficient; NfL, neurofilament light chain; p-tau181, phosphorylated tau 181; p-tau217, phosphorylated tau 217.

Supplementary Table 2. Model fit estimates and comparisons between the ACE model and the nested AE, CE and E sub-models.

| Model | ep | -2LL | df | Δ-2LL | Δdf | p | AIC |
| --- | --- | --- | --- | --- | --- | --- | --- |
| ACE | 34 | 6807.34 | 2710 | - | - | - | 6875.34 |
| AE | 24 | 6812.45 | 2720 | 5.11 | 10 | 0.884 | 6860.45 |
| CE | 24 | 6831.7 | 2720 | 24.35 | 10 | 0.007 | 6879.7 |
| E | 14 | 6957.28 | 2730 | 149.94 | 20 | <0.001 | 6985.28 |

Abbreviations: ∆-2LL, change in minus two log likelihood; ∆df, change in degrees of freedom; –2LL, minus two log likelihood; A, additive genetic effects; AIC, Akaike’s information criterion; C, common environmental effects; df, degrees of freedom; E, unique environmental effects; ep, number of estimated parameters.

Supplementary Table 3. Standardized estimates for additive genetic (A), common environmental (C) and unique environmental (E) variance components with 95% confidence intervals from the fitted AE model.

| Biomarker | A (95% CI) | C (95% CI) | E (95% CI) |
| --- | --- | --- | --- |
| p-tau181 | 0.13 (-0.41–0.71) | 0.24 (-0.28–0.66) | 0.63 (0.48–0.82) |
| p-tau217 | 0.48 (-0.06–1.04) | -0.03 (-0.53–0.41) | 0.54 (0.41–0.72) |
| NfL | 0.52 (0.09–0.96) | 0.07 (-0.33–0.41) | 0.42 (0.31–0.57) |
| GFAP | 0.50 (0.08–1.00) | 0.15 (-0.32–0.50) | 0.35 (0.26–0.48) |

Abbreviations: A, additive genetic effects; CI, confidence interval; E, unique environmental effects; GFAP, glial fibrillary acidic protein; NfL, neurofilament light chain; p-tau181, phosphorylated tau 181; p-tau217, phosphorylated tau 21

Supplementary Table 4. Model fit estimators in linear mixed-effects models. All models included sex, education and age as fixed effects, study site and family effects as random intercepts and total cognitive score as the outcome. Models 1–6 with biomarker concentrations as additional fixed effects were compared to Model 0.

| Model | Variables | ep | -2LL | df | Δ-2LL | Δdf | p | AIC | R2 | ΔR2 |
| --- | --- | --- | --- | --- | --- | --- | --- | --- | --- | --- |
| Model 0 |  | 7 | 1688.29 | 643 |  |  |  | 1702.29 | 0.19 |  |
| Model 1 | log(Aβ42) - log(Aβ40),  0.5 * (log(Aβ42) + log(Aβ40)) | 9 | 1685.55 | 641 | 2.74 | 2 | 0.748 | 1703.55 | 0.19 | 0 |
| Model 2 | p-tau181 | 8 | 1678.63 | 642 | 9.65 | 1 | 0.009 | 1694.63 | 0.2 | 0.01 |
| Model 3 | p-tau217 | 8 | 1659.52 | 642 | 28.77 | 1 | <0.001 | 1675.52 | 0.22 | 0.03 |
| Model 4 | NfL | 8 | 1682.93 | 642 | 5.36 | 1 | 0.076 | 1698.93 | 0.19 | 0.01 |
| Model 5 | GFAP | 8 | 1688.05 | 642 | 0.24 | 1 | >0.999 | 1704.05 | 0.19 | 0 |
| Model 6 | log(Aβ42) - log(Aβ40),  0.5 * (log(Aβ42) + log(Aβ40)), p-tau181, p-tau217,  NfL, GFAP | 13 | 1653.21 | 637 | 35.08 | 6 | <0.001 | 1679.21 | 0.23 | 0.04 |

Abbreviations: ∆-2LL, change in minus two log likelihood; ∆df, change in degrees of freedom; ΔR^2^, change in marginal R^2^ value, –2LL, minus two log likelihood; Aβ40, amyloid beta 40; Aβ42, amyloid beta 42; AIC, Akaike’s information criterion; df, degrees of freedom; ep, number of estimated parameters; GFAP, glial fibrillary acidic protein; NfL, neurofilament light chain; p-tau181, phosphorylated tau 181; p-tau217, phosphorylated tau 217.

Note: Adjusted p-values were calculated with the Benjamini–Yekutieli procedure to control the false discovery rate.
